# Long-Term Mortality Prediction with Pooled Cohort Equations: Implications for Lipid-Lowering Therapy

**DOI:** 10.1101/2024.04.05.24305406

**Authors:** Zhiyuan Ma, Lynn N. Moran, Jamshid Shirani

## Abstract

**Background:** Current guidelines recommend primary prevention statin therapy based on 10-year risk of atherosclerotic cardiovascular disease (ASCVD) assessed by the pooled cohort equations (PCE), while assessing effectiveness through low-density lipoprotein cholesterol levels. It remains uncertain whether 10-year ASCVD risk can accurately predict the mortality impact of lipid-lowering therapy.

**Methods:** A retrospective analysis of National Health and Nutrition Examination Survey (NHANES) (III and 1999-2008) linked to the National Death Index was conducted with propensity score matching. The study included 6647 adults without lipid-lowering therapy and 2484 with lipid-lowering therapy. Cox regression models and C statistic were used to assess the association between 10-year ASCVD risk and mortality.

**Results:** In the matched cohort with 4802 individuals with similar ASCVD risks, 10-year risk of ASCVD by PCE was comparable for predicating all-cause mortality at 10 years in the lipid-lowering therapy group (area under curve [AUC], 0.75; 95% CI 0.73-0.77) and without lipid-lowering therapy (0.74; 95% CI 0.71-0.77; *P* = 0.50). Similarly, PCE was comparable for predicting cardiovascular mortality at 10 years in both groups (AUC, 0.75; 95% CI 0.70-0.79 versus 0.77; 95% CI 0.73-0.80; *P* = 0.47). Lipid-lowering therapy was significantly associated with reduced all-cause mortality (adjusted hazard ratio [HR], 0.70; 95% CI 0.61-0.82; *P* < 0.01) and cardiovascular mortality (adjusted HR, 0.65; 95% CI, 0.51-0.83; *P* < 0.01), particularly in those with a 10-year ASCVD risk of 7.5% or higher.

**Conclusions:** PCE provides comparable predictions of mortality in individuals with and without lipid-lowering therapy. Moreover, individuals on lipid-lowering therapy exhibited lower all-cause and cardiovascular mortality compared to those without lipid-lowering therapy but with a similar 10-year risk of ASCVD. These findings suggest that mortality risk reduction can be assessed by 10-year ASCVD risk estimated by PCE for primary prevention both prior and after treatment.

---

Atherosclerotic cardiovascular disease (ASCVD) is a major cause of morbidity and mortality worldwide ^1,2^ and affects about 26 million people in the United States ^3,4^. The primary strategy to preventing ASCVD involves reducing the risk of developing ASCVD through lifestyle changes and the use of lipid-lowering therapy, such as statin therapy ^5^. Current guidelines recommend the use of the 10-year risk of ASCVD calculated from the pooled cohort equations (PCE) ^6^ to identify individuals at high risk of ASCVD who would benefit most from statin therapy ^7–10^. Statin therapy is recommended for adults aged 40 to 75 years with a 10-year risk of ASCVD of 7.5% or higher, with diabetes, or with low-density lipoprotein cholesterol (LDL-C) ≥ 190 mg/dL, and for those with clinical ASCVD ^8^. Many clinical trials have demonstrated that statin therapy reduces the risk of ASCVD and improve all-cause and cardiovascular mortality in individuals with varying baseline risks ^11–14^, particularly those with elevated LDL-C levels ^15^. In clinical practice, the effectiveness of statin therapy is typically assessed through monitoring of LDL-C levels. In general, reducing LDL-C levels by 1.0 mmol/L (about 39 mg/dL) is associated with about 25% reduction in the risk of CVD events ^16,17^.

However, the ability of the 10-year ASCVD risk derived from the PCE to predict mortality for individuals receiving lipid-lowering therapy and monitor the effectiveness of statin therapy has not been explored. In this study, we aimed to investigate the utility of the 10-year risk of ASCVD using the PCE for assessing mortality risk reduction in individuals without ASCVD, using data from the National Health and Nutrition Examination Survey (NHANES) (III and 1999-2008) linked to the National Death Index.

## Methods

### Study population

For this analysis, data from NHANES III and continuous NHANES 1999-2008 were included, in which civilian noninstitutionalized individuals in the United States were surveyed (https://www.cdc.gov/nchs/nhanes/index.htm). All individuals provided informed consent to participate in the NHANES study. The included NHANES study were approved by the NHANES Institutional Review Board and the NCHS Research Ethics Review Board. All survey data are deidentified and publicly available. This study followed the Strengthening the Reporting of Observational Studies in Epidemiology (STROBE) reporting guideline ^18^. Adults aged ≥ 40 and < 80 were included in the study. Individuals with a reported history of myocardial infarction, heart failure, coronary artery disease or stroke, those in whom 10-year ASCVD risk calculations were not feasible because of missing high-density lipoprotein cholesterol, total cholesterol (TC), or systolic blood pressure (SBP), or because of SBP < 90 mmHg or >200 mmHg, or TC <130 mg/dL or >320 mg/dL, and those with missing values for body mass index (BMI) and ineligible for mortality linkage were excluded. Individuals with missing self-reported responses to antihypertensive or lipid-lowering medications were included as those without therapy.

### Variable definitions

Hypertension was defined as systolic blood pressure ≥ 140 mmHg or diastolic blood pressure ≥ 90 mmHg, or self-reported use of antihypertensive medication. Diabetes was defined as hemoglobin A_1c_ concentration ≥ 6.5% or self-reported diagnosis of diabetes. We calculated 10-year risk of ASCVD based on 2018 ACC/AHA guidelines^8^ by the PCE^6^.

### Outcome measures

The primary outcome was mortality from all causes with truncating follow-up at 10 years. The secondary outcome was cardiovascular mortality. Mortality follow-up data were obtained from the Linked Mortality Files, which were collected from death certificate records from the National Death Index through December 31, 2019 using probabilistic linkage algorithm. Underlying Cause of Death Recodes (UCOD_113) in the Linked Mortality Files was created to assist mortality analyses using *International Classification of Diseases, 9^th^ Revision (ICD-9)* and *10th Revision (ICD-10) codes*. Death from cardiovascular diseases, which were derived from UCOD_113 codes (054-068 and 070) consisted of deaths from diseases of heart (I00-I09, I11, I13, I20-I51), including ischemic heart disease (I20 through I25), heart failure (I50) and essential hypertensive heart disease (I11 through I13), and cerebrovascular disease (I60 through I69).

### Statistical analysis

The discriminatory power of the risk of mortality as a function of 10-year ASCVD risk with or without lipid-lowering therapy was assessed by the area under receiver operating characteristic curve (AUC) (C statistic). The AUCs of the models with or without lipid-lowering therapy were compared for statistical differences with the use of bootstrapping.

To account for the potential confounders and to ensure the robustness of our results, propensity score matching was used based on variables including age, BMI, gender, race, education, 10-year ASCVD risk. Propensity score was estimated by a multivariable logistic regression model. The matched cohort was created at a 1:1 ratio with a caliper size of 0.05. The balance between covariates was evaluated by estimating standardized mean differences (SMD). SMD < 0.1 is considered a negligible group imbalance.

The Kaplan-Meier Method was used to estimate the cumulative event rate, and competing risks were modeled for cardiovascular mortality and death from other causes for the secondary outcome^19^. Individuals were categorized based on their 10-year ASCVD risk according to the 2018 ACC/AHA guidelines (< 5%, ≥ 5% to 7.4%, ≥ 7.5% to 19.9%, and ≥ 20%) for subgroup analysis. The risk of mortality for each ASCVD risk group with or without lipid-lowering therapy was estimated, adjusting for age, race, and BMI, with the <5% group as the reference. Cox proportional hazards models were used to assess the mortality benefit of lipid-lowering therapy versus control, with and without adjustment for age, race, hypertension, diabetes, smoking, and 10-year ASCVD risk. The unadjusted Cox regression was used to compare the treatment effect between each category, and the interaction between lipid-lowering therapy status and 10-year ASCVD risk was tested using interaction terms in the unadjusted Cox regression. Sensitivity analyses were conducted employing survey weights for Cox regression or utilizing individuals without lipid-lowering therapy from NHANES 1999-2008 as controls for propensity score matching.

Data were presented as mean and 95% confidence interval (CI) or median and interquartile range (IQR) for continuous variables. For differences in continuous variables, Student’s t tests were used in two groups for comparison. Categorical variables were expressed as percentage (%) and weights for examination and interview portions of survey were applied to account for the complex multistage probability-sampling design, survey non-response, and oversampling. For the comparison of categorical variables, χ2 tests were used. The risk of outcome was calculated as hazard ratios (HR) and 95% confidence intervals (CI). *P* values < 0.05 were considered to be statistically significant. All analyses were conducted with R (version 4.1.2)

## Results

A total of 6647 individuals without lipid-lowering therapy (47.4% men, median age 56 years [interquartile range, 46-66]) from NHANES III and 2484 with lipid-lowering therapy (47.1% men, median age 62 years [interquartile range, 54-70]) were matched based on similar 10-year risk of ASCVD, resulting in a cohort of 4802 individuals. In addition, a total of 8363 individuals without lipid-lowering therapy (49.2% men, median age 54 years [interquartile range, 46-64]) from NHANES 1999-2008 were used for sensitivity analysis. Individuals receiving lipid-lowering therapy had higher rate of hypertension (64.3% vs. 47.3%, *P* < .01) and diabetes (27.0% vs. 16.5%, *P* < .01) and lower rate of smoker (14.5% vs. 18.0%, *P* < .01), compared with those not receiving lipid-lowering therapy. The lipid-lowering therapy group had a lower 10-year all-cause mortality rate (13.4%, 321 of 2401) and cardiovascular mortality rate (4.4%, 106 of 2401), compared with those without lipid-lowering therapy (19.0%, 456 of 2401 and 6.9%, 165 of 2401, respectively). Moreover, a significant increase in lipid-lowering therapy was noted in the USA from the period 1984-1994 to the early 2000s (Supplementary figure 1). The flow diagram of patient selections was shown in Figure 1. The baseline characteristics of cohorts from NHANES III and 1999-2008 were summarized in Table 1.

**Figure 1.**
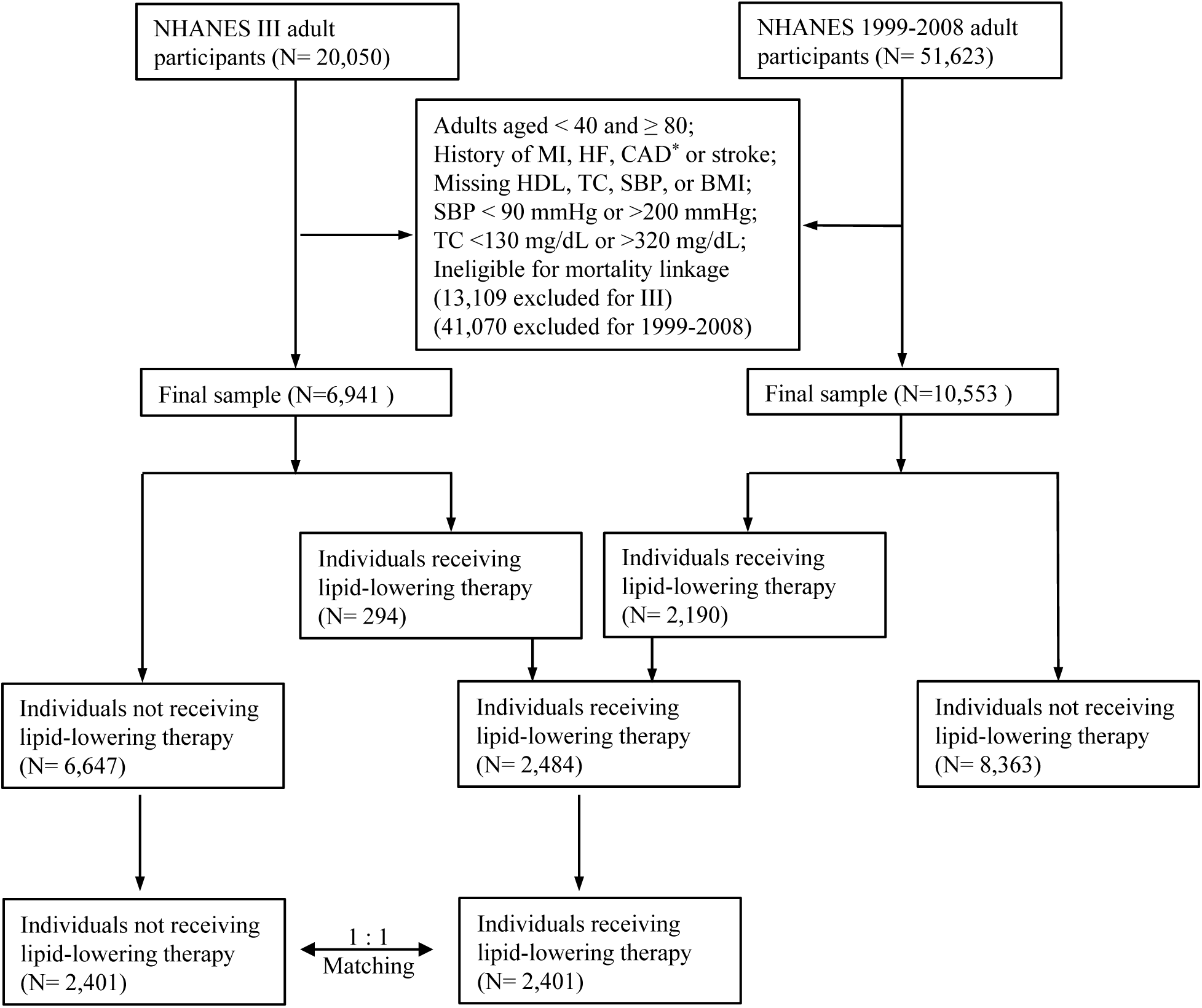
Flowchart illustration of the study cohorts. BMI, body mass index; CAD, coronary artery disease; HDL, high-density lipoprotein cholesterol; HF, heart failure; MI, myocardial infarction; SBP, systolic blood pressure; TC, total cholesterol. * CAD was not specified in NHANES III.

**Table 1.** Baseline characteristics of individuals by lipid-lowering therapy status.

|  | NHANESIII |  | NHANES1999-2008 |  | After Match<sup>a</sup> |  |  |
| --- | --- | --- | --- | --- | --- | --- | --- |
| Characteristic | Untreated (Control) | Lipid-lowering therapy | Untreated (Control) | Lipid-lowering therapy | Untreated (Control) | Lipid-lowering therapy | *P* Value |
| No. in sample | 6647 | 294 | 8363 | 2190 | 2401 | 2401 |  |
| Age, median [IQR] | 56 [46, 66] | 64 [57, 70] | 54 [46, 64] | 62 [53, 70] | 63 [53, 71] | 62 [53, 69] | 0.074 |
| Male, No. (weighted %) | 3150 (46.6) | 122 (44.9) | 4111 (46.7) | 1048 (49.0) | 1105 (46.0) | 1130 (47.1) | 0.487 |
| Race, No. (weighted %) |  |  |  |  |  |  | 0.932 |
| Mexican American | 1630 (3.6) | 51 (2.0) | 1847 (5.8) | 405 (4.3) | 439 (18.3) | 453 (18.9) |  |
| Non-Hispanic Black | 1677 (9.0) | 57 (5.8) | 1677 (9.8) | 377 (7.5) | 427 (17.8) | 430 (17.9) |  |
| Non-Hispanic White | 3070 (80.8) | 177 (85.1) | 4082 (75.7) | 1203 (79.1) | 1359 (56.6) | 1338 (55.7) |  |
| Other | 270 (6.6) | 9 (7.0) | 757 (8.7) | 205 (9.2) | 176 (7.3) | 180 (7.5) |  |
| BMI, mean (SD) | 27.9 (5.7) | 28.1 (4.7) | 28.7 (6.2) | 30.0 (5.9) | 29.5 (6.30) | 29.5 (5.49) | 0.712 |
| Education, No. (weighted %) |  |  |  |  |  |  | 0.544<sup>b</sup> |
| <9 years | 1827 (12.9) | 66 (12.7) | 1346 (6.6) | 363 (7.6) | 440 (18.3) | 422 (17.6) |  |
| >15 years | 945 (23.2) | 47 (24.5) | 1728 (28.1) | 438 (26.7) | 433 (18.0) | 460 (19.2) |  |
| 9-15 years | 3875 (63.8) | 181 (62.8) | 5282 (65.3) | 1388 (65.7) | 1528 (63.6) | 1519 (63.3) |  |
| Unknown | 0 (0.0) | 0 (0.0) | 7 (0.0) | 1 (0.0) | 0 (0.0) | 0 (0.0) |  |
| Hypertension, No. (weighted %) | 2682 (33.5) | 177 (55.5) | 3417 (34.8) | 1427 (61.5) | 1136 (47.3) | 1544 (64.3) | <0.001 |
| Diabetes, No. (weighted %) | 959 (9.1) | 62 (16.6) | 1016 (7.7) | 616 (22.2) | 395 (16.5) | 649 (27.0) | <0.001 |
| Currently smoking, No. (weighted %) | 1672 (24.4) | 41 (11.5) | 1621 (19.5) | 311 (15.3) | 432 (18.0) | 347 (14.5) | 0.001 |
| 10-year ASCVD risk, mean (SD) | 11.9 (12.1) | 17.2 (13.0) | 9.9 (11.2) | 14.9 (12.5) | 15.3 (13.0) | 15.0 (12.5) | 0.538 |
| 10-year ASCVD risk group, No. (weighted %) |  |  |  |  |  |  | <0.001 |
| Under 5% | 2530 (48.2) | 47 (26.0) | 3921 (58.6) | 510 (32.4) | 635 (26.4) | 551 (22.9) |  |
| 5-7.4% | 731 (11.6) | 32 (13.4) | 956 (11.6) | 261 (14.3) | 223 (9.3) | 287 (12.0) |  |
| 7.5-20% | 2013 (26.9) | 115 (32.6) | 2171 (20.6) | 813 (34.3) | 816 (34.0) | 892 (37.2) |  |
| 20% and over | 1373 (14.4) | 100 (28.0) | 1315 (9.2) | 606 (19.1) | 727 (30.3) | 671 (27.9) |  |
| Use of lipid-lowering therapy, weighted % | 4.5 |  | 19.7 |  | NA |  |  |

Abbreviations: ASCVD, atherosclerotic cardiovascular disease; BMI, body mass index; IQR, interquartile range.
<sup>a</sup> Percentage is derived from the cohort after match. <sup>b</sup> excluding the category of 'Unknown'.

### All-cause mortality

The incidence rate of all-cause mortality increased with increasing ASCVD risk category (Supplementary figure 2A and 2C). Cox proportional hazards analyses, using 10-year ASCVD risk < 5% as the reference, revealed significant associations with all-cause mortality for ASCVD risk ≥ 5% to 7.4% (adjusted HR, 1.74; 95% CI, 1.26-2.39; *P* < .01), ≥ 7.5% to 19.9% (adjusted HR, 2.02; 95% CI, 1.54-2.65; *P* < .01), and ≥ 20% (adjusted HR, 3.73; 95% CI, 2.75-5.05; *P* < .01) in the control group (Supplementary figure 2A and 3A). Similarly, individuals with lipid-lowering therapy had an increased for ASCVD risk ≥ 5% to 7.4%, ≥ 7.5% to 19.9%, and ≥ 20% with HRs of 1.24 (95% CI, 0.61-2.52; *P* = 0.50), 2.54 (95% CI, 1.48-4.37; *P* < .01), and 4.67 (95% CI, 2.60-8.37; *P* < .01), respectively (Supplementary figure 2C and 3B).

In the propensity score-matched cohort, co-variables, including age, BMI, gender, race, education and 10-year ASCVD risk, were well balanced (Figure 2A). The AUCs for 10-year ASCVD risk by PCE in control and lipid-lowering therapy groups for predicting all-cause mortality were similar (0.74; 95% CI, 0.71-0.77 versus 0.75; 95% CI, 0.73-0.77; *P* = .50) (Figure 2B). Lipid-lowering therapy was significantly associated with decreased all-cause mortality compared to control in the unadjusted analysis (HR, 0.68; 95% CI, 0.59-0.79; *P* < .01) (Figure 3A and 4A). The risk reduction remained statistically significant after multivariable adjustment (adjusted HR, 0.70; 95% CI 0.61-0.82; *P* < .01) (Figure 3A). In the subgroup analysis, individuals with a 10-year risk of ASCVD of 20% and over and ≥ 7.5% to 19.9%, were associated with a significantly lower risk of all-cause mortality with HRs of 0.66 (95% CI, 0.55-0.80; *P* < .01) and 0.69 (95% CI, 0.54-0.88; *P* < .01) (Figure 3A and 4B). There was no significant interaction with 10-year ASCVD risk seen for lipid-lowering therapy status (*P* = .99 for interaction) (Figure 3A).

**Figure 2.**
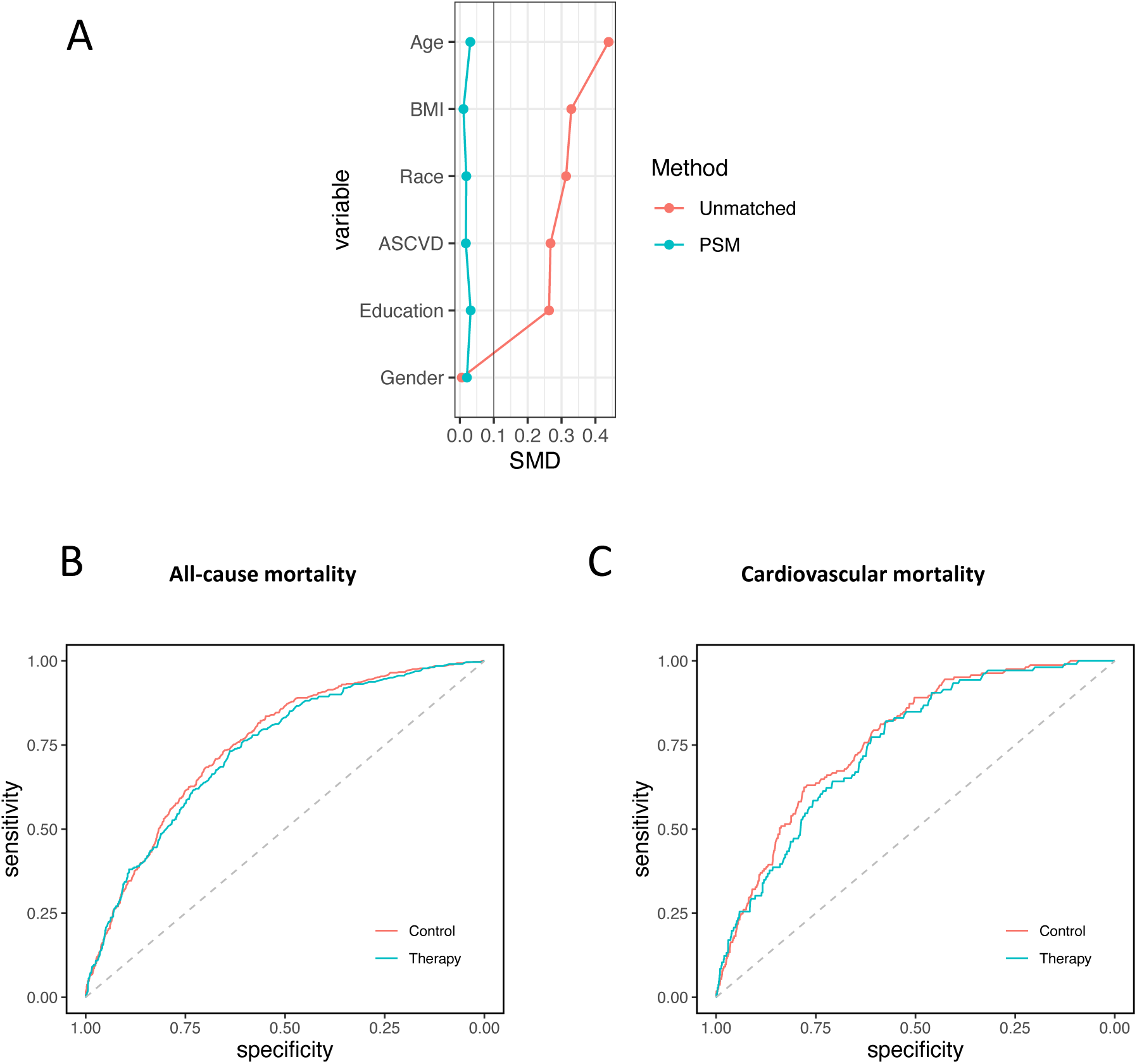
ROC analysis of 10-year ASCVD risk to predict mortality in the matched cohort. (A) Changes in standardized mean difference (SMD) before and after matching. (B) Predicting all-cause mortality in untreated control and lipid-lowering therapy groups (AUC [95% CI]: 0.74 [0.71-0.77] versus 0.75[ 0.73-0.77]; *P* = .50). (C) Predicting cardiovascular mortality in untreated control and lipid-lowering therapy groups (AUC [95% CI]: 0.77 [0.73-0.80] versus 0.75 [0.70-0.79]; P = .47). PSM: propensity score matching.

**Figure 3.**
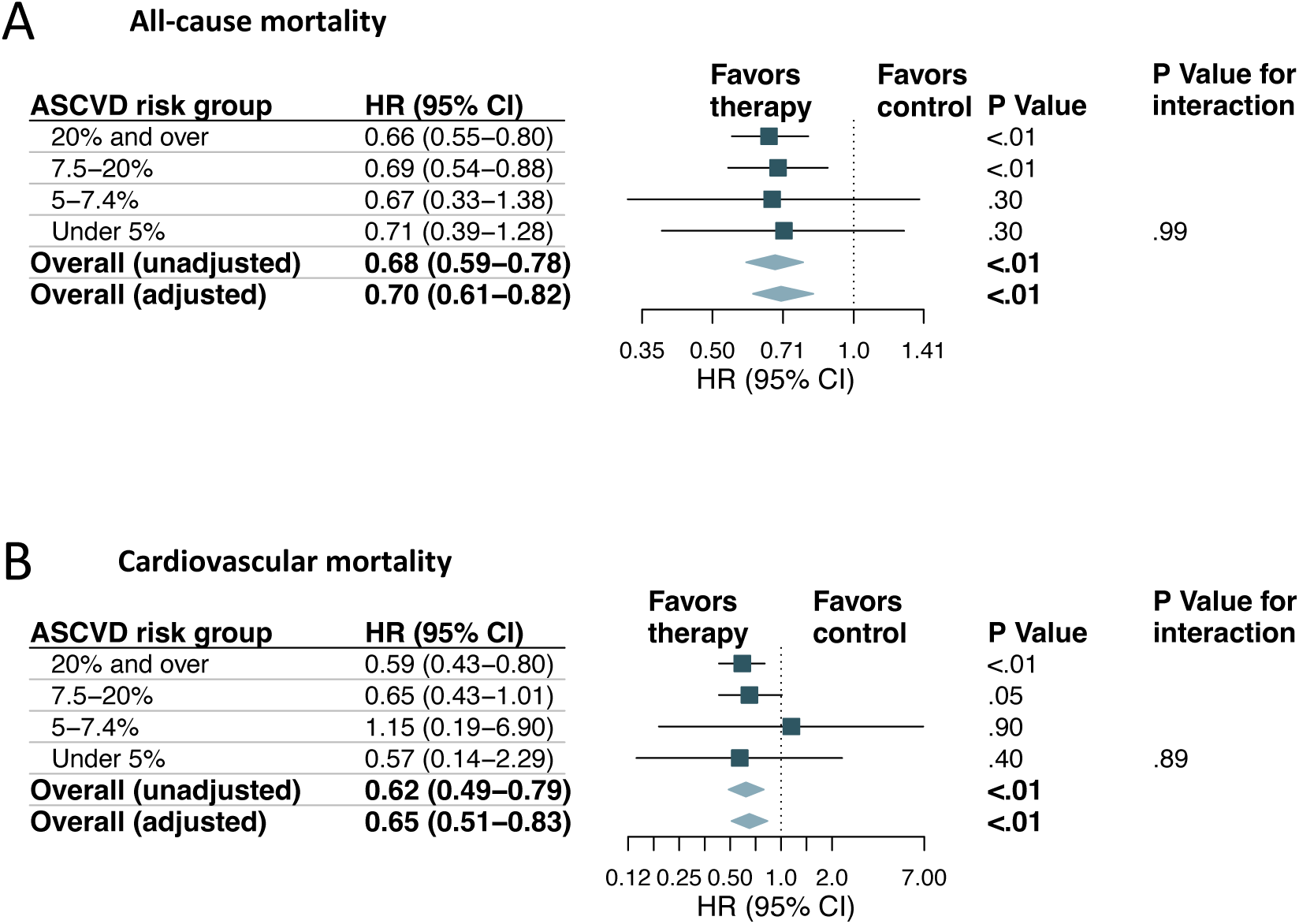
Analysis of mortality outcomes in the matched cohort. (A) Hazard ratios and 95% confidence intervals are shown for all-cause mortality in the total cohort and subgroups with increasing categories of 10-year ASCVD risk. (B) Hazard ratios and 95% confidence intervals are shown for cardiovascular mortality in the total cohort and subgroups with increasing categories of 10-year ASCVD risk.

**Figure 4.**
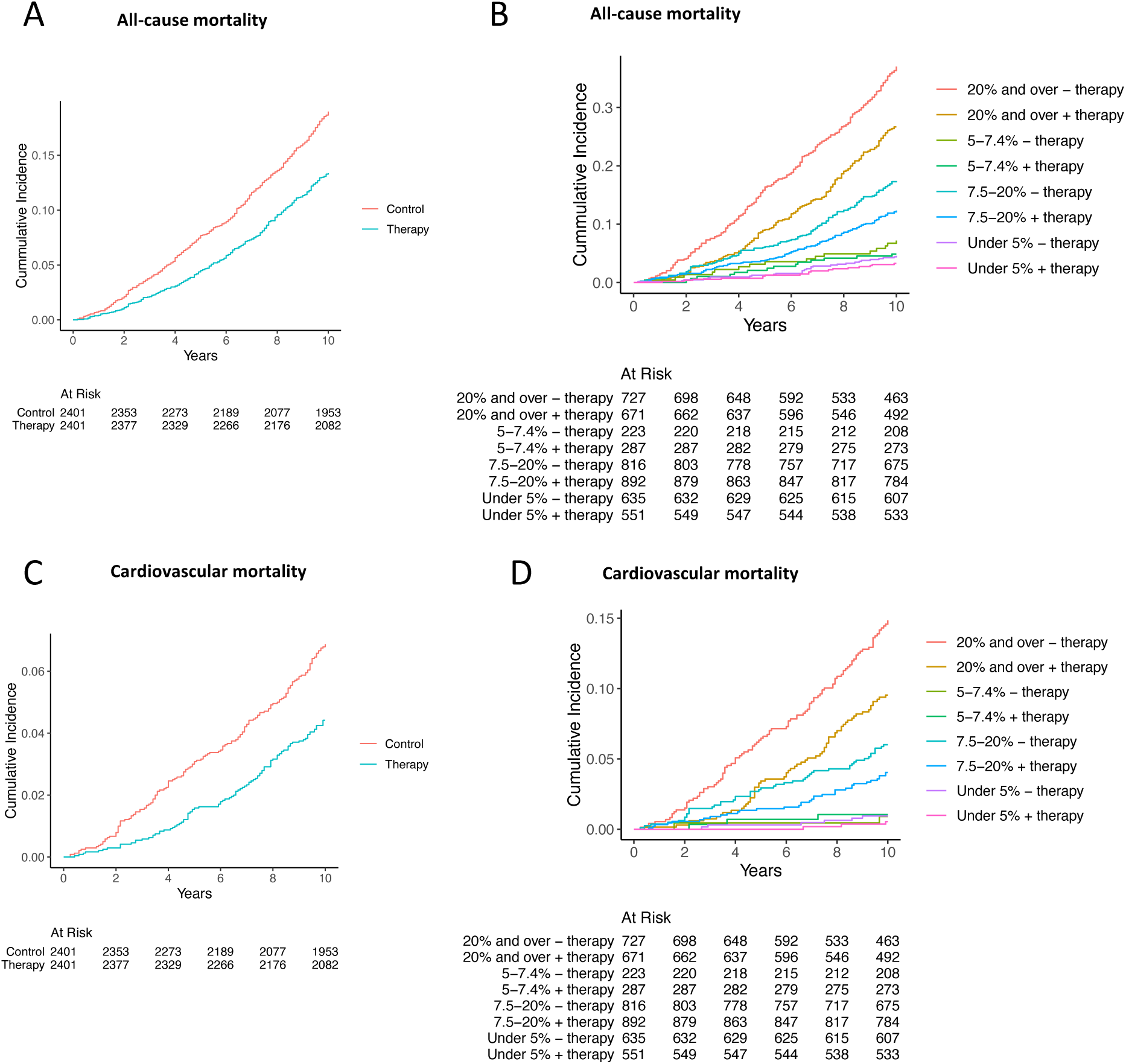
Cumulative event rate of primary and secondary outcomes in the matched cohort. (A) All-cause mortality in individuals who were receiving lipid-lowering therapy, compared to those who were not. (B) All-cause mortality in subgroups with increasing categories of 10-year ASCVD risk. (C) Cardiovascular mortality in individuals who were receiving lipid-lowering therapy, compared to those who were not.

In the unmatched cohort, the AUC for 10-year ASCVD risk by PCE in control for predicting all-cause mortality was slightly higher than that for lipid-lowering therapy groups (0.78; 95% CI, 0.76-0.79 versus 0.74; 95% CI, 0.71-0.76; *P* = .02) (Supplementary figure 4A). Additionally, lipid-lowering therapy was found to be significantly associated with reduced all-cause mortality in the unadjusted analysis when compared to the control group (HR, 0.88; 95% CI 0.78-0.99; *P* = .04) (Supplementary figure 5A and 6A). The risk reduction remained statistically significant after multivariable adjustment (adjusted HR, 0.70; 95% CI 0.61-0.79; *P* < .01) (Supplementary figure 5A). Furthermore, in the subgroup analysis, individuals with a 10-year risk of ASCVD of ≥ 20%, ≥ 7.5% to 19.9%, and ≥ 5% to 7.4% were associated with significantly lower all-cause mortality with HRs of 0.65 (95% CI, 0.55-0.77; *P* < .01), 0.75 (95% CI, 0.60-0.93; *P* < .01), and 0.51 (95% CI, 0.29-0.91; *P* = .02), respectively (Supplementary figure 5A and 6B). Notably, no significant interaction was observed with 10-year ASCVD risk for lipid-lowering therapy status (*P* = .43 for interaction) (Supplementary figure 5A).

### Cardiovascular mortality

The incidence rate of cardiovascular mortality rose with increasing ASCVD risk category (Supplementary figure 2B and 2D). In Cox proportional hazards analyses, using 10-year ASCVD risk < 5% as the reference, there was significant associations with cardiovascular mortality for ASCVD risk ≥ 5% to 7.4% (adjusted HR, 2.07; 95% CI, 1.00-4.28; P = .05), ≥ 7.5% to 19.9% (adjusted HR, 3.46; 95% CI, 1.92-6.24; P < .01), and ≥ 20% (adjusted HR, 7.46; 95% CI, 3.96-14.10; P < .01) in the control group (Supplementary figure 2B and 3A). Similarly, increased risk of cardiovascular mortality was observed in individuals with lipid-lowering therapy for the ASCVD risk ≥ 5% to 7.4%, ≥ 7.5% to 19.9%, and ≥ 20% with HRs of 1.64 (95% CI, 0.33-8.19; P = 0.5), 5.45 (95% CI, 1.59-18.7; P < .01), and 11.6 (95% CI, 3.19-42.00; P < .01), respectively (Supplementary figure 2D and 3B).

In the propensity score-matched cohort, the AUCs for 10-year ASCVD risk by PCE in control and lipid-lowering therapy groups for predicting cardiovascular mortality were similar (0.77; 95% CI, 0.73-0.80 versus 0.75; 95% CI, 0.70-0.79; P = .47) (Figure 2C). Compared to control, lipid-lowering therapy was significantly associated with decreased cardiovascular mortality in the unadjusted analysis (HR, 0.62; 95% CI 0.49-0.79; P < .01) (Figure 3B and 4C). The risk reduction remained statistically significant after multivariable adjustment (adjusted HR, 0.65; 95% CI 0.51-0.83; P < .01) (Figure 3B). In the subgroup analysis, individuals with a 10-year risk of ASCVD of 20% and over, were associated with a significantly lower risk for cardiovascular mortality with a HR of 0.59 (95% CI, 0.43-0.80; P < .01) (Figure 3B and 4D). Test for interaction between lipid-lowering therapy status and 10-year ASCVD risk was not significant (*P* = .89 for interaction) (Figure 3B).

In the unmatched cohort, the AUC for 10-year ASCVD risk in the control group for predicting cardiovascular mortality was slightly higher compared to that for lipid-lowering therapy groups (0.80; 95% CI, 0.78-0.82 versus 0.75; 95% CI, 0.70-0.79; P = .02) (Supplementary figure 4B). In the unadjusted analysis, lipid-lowering therapy was not significantly associated with decreased cardiovascular mortality compared to the control group (HR, 0.88; 95% CI 0.71-1.09; P = .30) (Supplementary 5B and 6C). However, after multivariable adjustment, the risk reduction was statistically significant (adjusted HR, 0.66; 95% CI 0.53-0.83; P < .01) (Supplementary figure 5B). In the subgroup analysis, individuals with a 10-year risk of ASCVD of ≥ 20% were associated with a significantly lower cardiovascular mortality with a HR of 0.60 (95% CI, 0.46-0.80; P < .01) (Supplementary figure 5B and 6D). There was no significant interaction with 10-year ASCVD risk observed for lipid-lowering therapy status (P = .72 for interaction) (Supplementary figure 5B).

### Sensitivity analysis

To investigate whether the beneficial effect of lipid-lowering therapy on mortality results from control from early NHANES cycles, individuals without lipid-lowering therapy from NHANES 1999-2008 were used as controls for propensity score matching. In the matched cohort with balanced covariates (Supplementary figure 7A), the AUCs for 10-year ASCVD risk by PCE in control and lipid-lowering therapy groups for predicting all-cause (0.73; 95% CI, 0.71-0.76 versus 0.74; 95% CI, 0.71-0.76; *P* = .80) and cardiovascular (0.76; 95% CI, 0.71-0.81 versus 0.75; 95% CI, 0.70-0.79; *P* = .67) mortality were similar (Supplementary figure 7A and 7B). Compared to control, lipid-lowering therapy was significantly associated with decreased all-cause mortality in both unadjusted (HR, 0.76; 95% CI 0.66-0.88; *P* < .01) and adjusted analyses (adjusted HR, 0.77; 95% CI 0.66-0.89; *P* < .01) (Supplementary figure 8A and 9A). However, the risk reduction for cardiovascular mortality was neither seen in the unadjusted analysis (HR, 1.19; 95% CI 0.90-1.57; *P* = .20), nor in the adjusted analysis (adjusted HR, 1.17; 95% CI 0.88-1.56; P = .30) (Supplementary figure 8B and 9C). In the subgroup analysis, individuals with a 10-year risk of ASCVD of 20% and over and ≥ 7.5% to 19.9%, were associated with a significantly lower risk for all-cause mortality with HRs of 0.78 (95% CI, 0.64-0.94; *P* = .01) and 0.65 (95% CI, 0.51-0.83; *P* < .01) (Supplementary figure 8A and 9B), but not cardiovascular mortality (Supplementary figure 8B and 9D). Furthermore, comparable outcomes were obtained in the matched cohort from NHANES 1999-2008, utilizing survey weights, to ascertain the potential impact of the intricate survey design on the mortality benefit associated with lipid-lowering therapy (Supplementary figures 10-12).

## Discussion

This study based on NHAES participants from period 1988-1994 and 1999-2008, found that the ability of 10-year ASCVD risk using the PCE to predict all-cause and cardiovascular mortality was comparable for individuals who were receiving lipid-lowering therapy and those who were not, after adjusted for the propensity to receive lipid-lowering therapy. Furthermore, there was an association between lipid-lowering therapy and the reduction of all-cause and cardiovascular mortality, with the greatest reductions for individuals with 10-year ASCVD risk of ≥ 7.5%.

In clinical practice, the 10-year risk of ASCVD calculated using the PCE has been used to identify individuals who are at high risk of ASCVD, but its prognostic value for lipid-lowering therapy has not been explored. It is crucial to use the same metric to assess the effectiveness of lipid-lowering therapy on all-cause and cardiovascular mortality for individuals before and after therapy. To determine the association of 10-year ASCVD risk and baseline mortality with minimal confounding from lipid-lowering therapy, eligible individuals without lipid-lowering therapy from NHANES III were included in this study, as a significant increase in lipid-lowering therapy was noted in the USA from the period 1984-1994 (NHANES III) to the early 2000s, which is in accordance with prior findings that the use of lipid-lowering therapy increased significantly after the early 2000s ^20^. For comparison, individuals with lipid-lowering therapy from the early 2000s were included. However, the limitations of this approach are discussed below.

Statins have been widely used as the primary lipid-lowering drugs. Several clinical trials and meta-analysis have demonstrated their beneficial effects on all-cause mortality and cardiovascular mortality in individuals with or without cardiovascular disease ^11,13–15,17,21–23^. Consistent with this, we found that lipid-lowering therapy was significantly associated with a 30% reduction in all-cause mortality and a 35% reduction in cardiovascular mortality in matched individuals with comparable 10-year risk of ASCVD. In the subgroup analysis, the greatest reductions were observed in individuals with 10-year ASCVD risk of ≥ 7.5%. Importantly, the all-mortality benefit remained significant in the unmatched cohort and sensitivity analysis, providing further evidence of the effectiveness of lipid-lowering therapy in reducing all-cause mortality. However, the cardiovascular mortality benefit associated with lipid-lowering therapy was not significant in the sensitivity analysis where different controls from a later time period (NHANES 1999-2008) were used. This finding suggests that the results of the propensity score-matched analysis may be sensitive to the choice of control group and time period used for comparison. There are several possible explanations for the differences. One possible explanation is that the control group in NHANES III may differ in important ways from the control group in NHANES 1999-2008, such as in terms of comorbidities or disease severity that were not accounted for in the matching process. Another possible explanation is that individuals in NHANES 1999-2008 are more likely to receive lipid-lowering therapy during later years after their initial interview compared to those in NHANES III, which could potentially confound the observed cardiovascular benefits of lipid-lowering therapy. Nonetheless, our study found that individuals who were taking lipid-lowering therapy had lower all-cause mortality rates and at least comparable or better cardiovascular mortality rates compared to those who were not taking lipid-lowering therapy but had similar 10-year risk of ASCVD. These results provide a basis for assessing mortality risk reduction using 10-year ASCVD risk by PCE as a key tool in individuals before and after lipid-lowering treatment, in addition to LDL-C levels.

It’s crucial to recognize that the PCE were derived to gauge the 10-year risk of clinical ASCVD, encompassing conditions like acute coronary syndrome, a history of myocardial infarction, stable or unstable angina, coronary or other arterial revascularization, stroke, transient ischemic attack, or peripheral artery disease, all stemming from atherosclerotic origins ^6,8^. While there have been reports indicating an overestimation of ASCVD relative risk by these equations ^24,25^, recent studies have demonstrated their robust performance within real-world community cohorts, irrespective of age, blood pressure, blood cholesterol levels, or the initiation of statin therapy during follow-up ^26^. These findings have further strengthened the utilization of PCE in assessing ASCVD risk. Our data corroborate these findings, demonstrating that the PCE consistently predict both all-cause and cardiovascular mortality regardless of lipid-lowering therapy status. This strengthens the case for employing the 10-year ASCVD risk estimated by PCE to monitor the relative reduction in mortality risk associated with lipid-lowering therapy.

The present study has several limitations. First, the lack of subsequent data on initiation and discontinuation of lipid-lowering therapy after the initial interview in NHANES could have confounded the observed benefits. Second, a considerable number of participants did not provide responses regarding their lipid-lowering therapy status in NHANES, which could introduce bias when assuming non-therapy status for analyses. However, similar conclusions were obtained when analyses were restricted to individuals who explicitly reported receiving lipid-lowering therapy or not (Data not shown). Third, the study did not document specific medications for lipid-lowering therapy, which may have included classes of medication beyond statins. Fourth, despite the use of propensity score matching and multiple variable adjustments to balance and control for potential confounding factors, residual confounders are likely to exist and could not be fully accounted for in this retrospective cohort study. Lastly, the sample size of the subgroup with borderline risk (5-7.4%) may have been too small to detect a statistically significant difference, indicating the need for further investigation on whether lipid-lowering therapy provides greater benefits to individuals with borderline ASCVD risk after treatment compared to those who do not receive such treatment.

In conclusion, the results of this study using NHANES data with a follow-up period of at least 10 years show that the 10-year risk of ASCVD calculated using the PCE has a comparable prognostic value for all-cause and cardiovascular mortality, irrespective of lipid-lowering therapy status. Additionally, our findings demonstrate that lipid-lowering therapy is associated with a reduction in both all-cause and cardiovascular mortality in individuals receiving treatment, particularly for those with a 10-year ASCVD risk of 7.5% or higher, compared to those not receiving treatment but with similar ASCVD risk. These results highlight the great potential of using the 10-year risk of ASCVD using the PCE for assessing mortality risk reduction in individuals before and after treatment for the prevention of ASCVD.

## Data Availability

All data produced in the present work are contained in the manuscript

## Acknowledgements

None

## References

1. DALYs GBD, Collaborators H. Global, regional, and national disability-adjusted life-years (DALYs) for 315 diseases and injuries and healthy life expectancy (HALE), 1990-2015: a systematic analysis for the Global Burden of Disease Study 2015. Lancet. 2016;388:1603–1658. doi: 10.1016/S0140-6736(16)31460-X

2. Mortality GBD, Causes of Death C. Global, regional, and national life expectancy, all-cause mortality, and cause-specific mortality for 249 causes of death, 1980-2015: a systematic analysis for the Global Burden of Disease Study 2015. Lancet. 2016;388:1459–1544. doi: 10.1016/S0140-6736(16)31012-1

3. Vasan RS, Enserro DM, Xanthakis V, Beiser AS, Seshadri S. Temporal Trends in the Remaining Lifetime Risk of Cardiovascular Disease Among Middle-Aged Adults Across 6 Decades: The Framingham Study. Circulation. 2022;145:1324–1338. doi: 10.1161/CIRCULATIONAHA.121.057889

4. Tsao CW, Aday AW, Almarzooq ZI, Anderson CAM, Arora P, Avery CL, Baker-Smith CM, Beaton AZ, Boehme AK, Buxton AE, et al. Heart Disease and Stroke Statistics-2023 Update: A Report From the American Heart Association. Circulation. 2023;147:e93–e621. doi: 10.1161/CIR.0000000000001123

5. Jaspers NEM, Blaha MJ, Matsushita K, van der Schouw YT, Wareham NJ, Khaw KT, Geisel MH, Lehmann N, Erbel R, Jockel KH, et al. Prediction of individualized lifetime benefit from cholesterol lowering, blood pressure lowering, antithrombotic therapy, and smoking cessation in apparently healthy people. Eur Heart J. 2020;41:1190–1199. doi: 10.1093/eurheartj/ehz239

6. Stone NJ, Robinson JG, Lichtenstein AH, Bairey Merz CN, Blum CB, Eckel RH, Goldberg AC, Gordon D, Levy D, Lloyd-Jones DM, et al. 2013 ACC/AHA guideline on the treatment of blood cholesterol to reduce atherosclerotic cardiovascular risk in adults: a report of the American College of Cardiology/American Heart Association Task Force on Practice Guidelines. Circulation. 2014;129:S1–45. doi: 10.1161/01.cir.0000437738.63853.7a

7. Grundy SM, Stone NJ, Bailey AL, Beam C, Birtcher KK, Blumenthal RS, Braun LT, de Ferranti S, Faiella-Tommasino J, Forman DE, et al. 2018 AHA/ACC/AACVPR/AAPA/ABC/ACPM/ADA/AGS/APhA/ASPC/NLA/PCNA Guideline on the Management of Blood Cholesterol: Executive Summary: A Report of the American College of Cardiology/American Heart Association Task Force on Clinical Practice Guidelines. J Am Coll Cardiol. 2019;73:3168–3209. doi: 10.1016/j.jacc.2018.11.002

8. Grundy SM, Stone NJ, Bailey AL, Beam C, Birtcher KK, Blumenthal RS, Braun LT, de Ferranti S, Faiella-Tommasino J, Forman DE, et al. 2018 AHA/ACC/AACVPR/AAPA/ABC/ACPM/ADA/AGS/APhA/ASPC/NLA/PCNA Guideline on the Management of Blood Cholesterol: A Report of the American College of Cardiology/American Heart Association Task Force on Clinical Practice Guidelines. Circulation. 2019;139:e1082–e1143. doi: 10.1161/CIR.0000000000000625

9. Grundy SM, Stone NJ, Guideline Writing Committee for the Cholesterol G. 2018 Cholesterol Clinical Practice Guidelines: Synopsis of the 2018 American Heart Association/American College of Cardiology/Multisociety Cholesterol Guideline. Ann Intern Med. 2019;170:779–783. doi: 10.7326/M19-0365

10. Force USPST, Mangione CM, Barry MJ, Nicholson WK, Cabana M, Chelmow D, Coker TR, Davis EM, Donahue KE, Jaen CR, et al. Statin Use for the Primary Prevention of Cardiovascular Disease in Adults: US Preventive Services Task Force Recommendation Statement. JAMA. 2022;328:746–753. doi: 10.1001/jama.2022.13044

11. Randomised trial of cholesterol lowering in 4444 patients with coronary heart disease: the Scandinavian Simvastatin Survival Study (4S). Lancet. 1994;344:1383–1389.

12. Ridker PM, Danielson E, Fonseca FA, Genest J, Gotto AM, Jr., Kastelein JJ, Koenig W, Libby P, Lorenzatti AJ, MacFadyen JG, et al. Rosuvastatin to prevent vascular events in men and women with elevated C-reactive protein. N Engl J Med. 2008;359:2195–2207. doi: 10.1056/NEJMoa0807646

13. Sacks FM, Pfeffer MA, Moye LA, Rouleau JL, Rutherford JD, Cole TG, Brown L, Warnica JW, Arnold JM, Wun CC, et al. The effect of pravastatin on coronary events after myocardial infarction in patients with average cholesterol levels. Cholesterol and Recurrent Events Trial investigators. N Engl J Med. 1996;335:1001–1009. doi: 10.1056/NEJM199610033351401

14. Sever PS, Dahlof B, Poulter NR, Wedel H, Beevers G, Caulfield M, Collins R, Kjeldsen SE, Kristinsson A, McInnes GT, et al. Prevention of coronary and stroke events with atorvastatin in hypertensive patients who have average or lower-than-average cholesterol concentrations, in the Anglo-Scandinavian Cardiac Outcomes Trial--Lipid Lowering Arm (ASCOT-LLA): a multicentre randomised controlled trial. Lancet. 2003;361:1149–1158. doi: 10.1016/S0140-6736(03)12948-0

15. Shepherd J, Cobbe SM, Ford I, Isles CG, Lorimer AR, MacFarlane PW, McKillop JH, Packard CJ. Prevention of coronary heart disease with pravastatin in men with hypercholesterolemia. West of Scotland Coronary Prevention Study Group. N Engl J Med. 1995;333:1301–1307. doi: 10.1056/NEJM199511163332001

16. Ference BA, Ginsberg HN, Graham I, Ray KK, Packard CJ, Bruckert E, Hegele RA, Krauss RM, Raal FJ, Schunkert H, et al. Low-density lipoproteins cause atherosclerotic cardiovascular disease. 1. Evidence from genetic, epidemiologic, and clinical studies. A consensus statement from the European Atherosclerosis Society Consensus Panel. Eur Heart J. 2017;38:2459–2472. doi: 10.1093/eurheartj/ehx144

17. Cholesterol Treatment Trialists C, Baigent C, Blackwell L, Emberson J, Holland LE, Reith C, Bhala N, Peto R, Barnes EH, Keech A, et al. Efficacy and safety of more intensive lowering of LDL cholesterol: a meta-analysis of data from 170,000 participants in 26 randomised trials. Lancet. 2010;376:1670–1681. doi: 10.1016/S0140-6736(10)61350-5

18. von Elm E, Altman DG, Egger M, Pocock SJ, Gotzsche PC, Vandenbroucke JP, Initiative S. The Strengthening the Reporting of Observational Studies in Epidemiology (STROBE) statement: guidelines for reporting observational studies. Lancet. 2007;370:1453–1457. doi: 10.1016/S0140-6736(07)61602-X

19. Austin PC, Lee DS, Fine JP. Introduction to the Analysis of Survival Data in the Presence of Competing Risks. Circulation. 2016;133:601–609. doi: 10.1161/CIRCULATIONAHA.115.017719

20. Ma J, Sehgal NL, Ayanian JZ, Stafford RS. National trends in statin use by coronary heart disease risk category. PLoS Med. 2005;2:e123. doi: 10.1371/journal.pmed.0020123

21. Downs JR, Clearfield M, Weis S, Whitney E, Shapiro DR, Beere PA, Langendorfer A, Stein EA, Kruyer W, Gotto AM, Jr. Primary prevention of acute coronary events with lovastatin in men and women with average cholesterol levels: results of AFCAPS/TexCAPS. Air Force/Texas Coronary Atherosclerosis Prevention Study. JAMA. 1998;279:1615–1622. doi: 10.1001/jama.279.20.1615

22. Yusuf S, Bosch J, Dagenais G, Zhu J, Xavier D, Liu L, Pais P, Lopez-Jaramillo P, Leiter LA, Dans A, et al. Cholesterol Lowering in Intermediate-Risk Persons without Cardiovascular Disease. N Engl J Med. 2016;374:2021–2031. doi: 10.1056/NEJMoa1600176

23. Nowak MM, Niemczyk M, Florczyk M, Kurzyna M, Paczek L. Effect of Statins on All-Cause Mortality in Adults: A Systematic Review and Meta-Analysis of Propensity Score-Matched Studies. J Clin Med. 2022;11. doi: 10.3390/jcm11195643

24. Cook NR, Ridker PM. Calibration of the Pooled Cohort Equations for Atherosclerotic Cardiovascular Disease: An Update. Ann Intern Med. 2016;165:786–794. doi: 10.7326/M16-1739

25. Pennells L, Kaptoge S, Wood A, Sweeting M, Zhao X, White I, Burgess S, Willeit P, Bolton T, Moons KGM, et al. Equalization of four cardiovascular risk algorithms after systematic recalibration: individual-participant meta-analysis of 86 prospective studies. Eur Heart J. 2019;40:621–631. doi: 10.1093/eurheartj/ehy653

26. Medina-Inojosa JR, Somers VK, Garcia M, Thomas RJ, Allison T, Chaudry R, Wood-Wentz CM, Bailey KR, Mulvagh SL, Lopez-Jimenez F. Performance of the ACC/AHA Pooled Cohort Cardiovascular Risk Equations in Clinical Practice. J Am Coll Cardiol. 2023;82:1499–1508. doi: 10.1016/j.jacc.2023.07.018

