## Supplemental Data for "Long-Term Mortality Prediction with Pooled Cohort Equations: Implications for Lipid-Lowering Therapy"

Insights from the National Health and Nutrition Examination Survey

Zhiyuan Ma, MD, PhD,[a, b] Lynn N. Moran, DO,[a] Jamshid Shirani, MD[a]

From the [a]Departments of Cardiology and [b]Research and Innovation, St. Luke's University Health Network, Bethlehem, PA

Address for correspondence: Zhiyuan Ma or Jamshid Shirani, St. Luke's University Health Network, 801 Ostrum Street, Bethlehem, PA 18015. Telephone: 484-526-4011; FAX: 484-526-4010; (ZM); (JS).

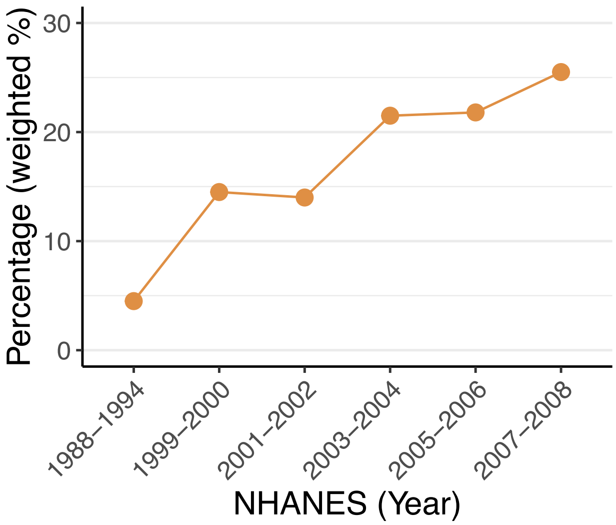


Supplementary figure 1. The weighted percentage of lipid-lowering therapy in the NHANES from 1988 to 2008.

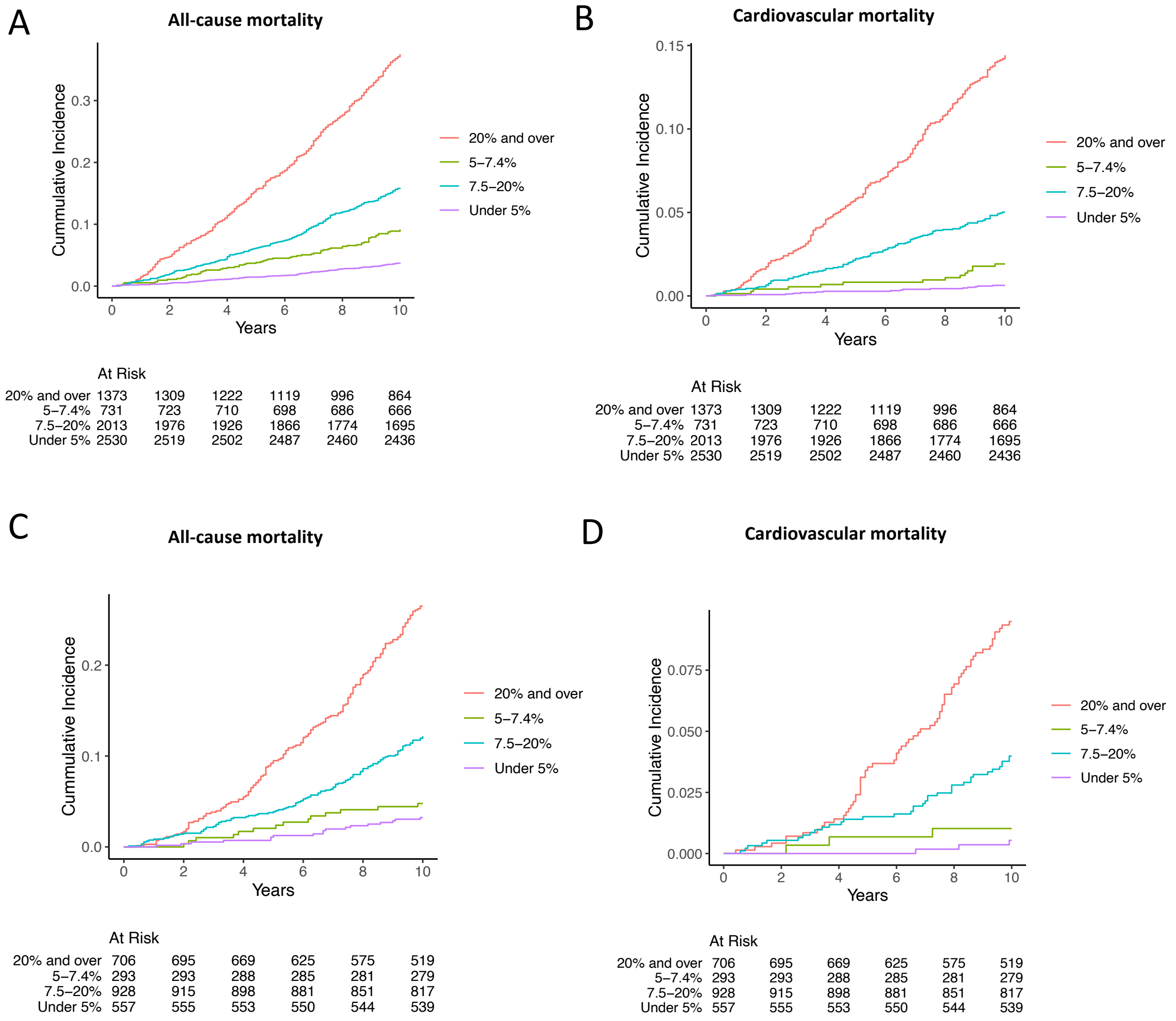


Supplementary figure 2. Cumulative incidence of each 10-year ASCVD risk group. (A) All-cause mortality in individuals who were not receiving lipid-lowering therapy. (B) Cardiovascular mortality in individuals who were not receiving lipid-lowering therapy. (C) All-cause mortality in individuals who were receiving lipid-lowering therapy. (D) Cardiovascular mortality in individuals who were receiving lipid-lowering therapy.

### A Subjects without lipid-lowering therapy

| Outcome | HR (95% CI) | P Value |
|---|---|---|
| **All–cause mortality** | | |
| 20% and over | 3.73 (2.75–5.05) | <.01 |
| 7.5–20% | 2.02 (1.54–2.65) | <.01 |
| 5–7.4% | 1.74 (1.26–2.39) | <.01 |
| Under 5% (reference) | – | |
| **Cardiovascular mortality** | | |
| 20% and over | 7.46 (3.96–14.10) | <.01 |
| 7.5–20% | 3.46 (1.92–6.24) | <.01 |
| 5–7.4% | 2.07 (1.00–4.28) | .05 |
| Under 5% (reference) | – | |

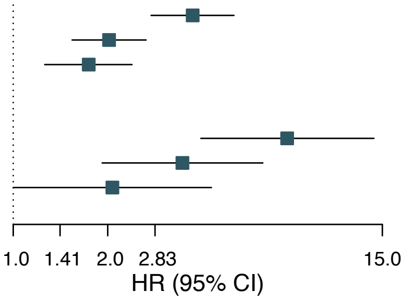


### B Subjects with lipid-lowering therapy

| Outcome | HR (95% CI) | P Value |
|---|---|---|
| **All–cause mortality** | | |
| 20% and over | 4.67 (2.60–8.37) | <.01 |
| 7.5–20% | 2.54 (1.48–4.37) | <.01 |
| 5–7.4% | 1.24 (0.61–2.52) | .50 |
| Under 5% (reference) | – | |
| **Cardiovascular mortality** | | |
| 20% and over | 11.60 (3.19–42.00) | <.01 |
| 7.5–20% | 5.45 (1.59–18.7) | <.01 |
| 5–7.4% | 1.64 (0.33–8.19) | .50 |
| Under 5% (reference) | – | |

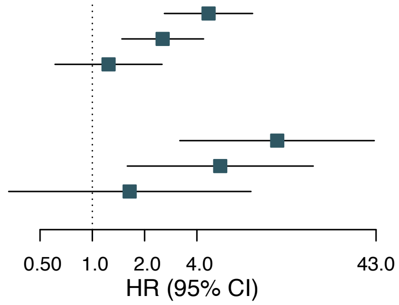


Supplementary figure 3. Hazard ratios for each 10-year ASCVD risk group without (A) or with (B) lipid-lowering therapy using ASCVD risk < 5% group as the reference group.

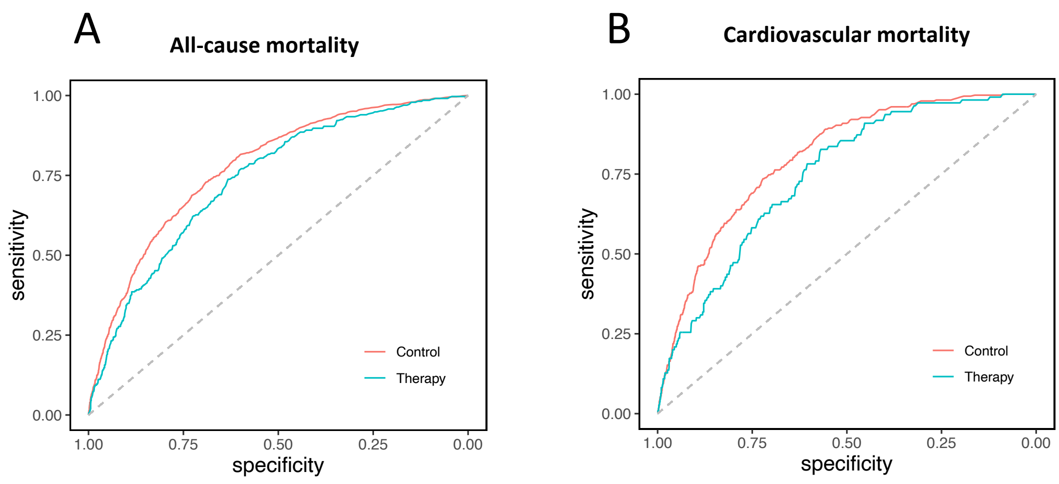


Supplementary figure 4. ROC analysis of 10-year ASCVD risk to predict mortality in the unmatched cohort. (A) Predicting all-cause mortality in untreated control and lipid-lowering therapy groups (AUC [95% CI]: 0.78 [0.76-0.79] versus 0.74 [ 0.71-0.76]; P = .02). (B) Predicting cardiovascular mortality in untreated control and lipid-lowering therapy groups (AUC [95% CI]: 0.80 [0.78-0.82] versus 0.75 [0.70-0.79]; P = .02).

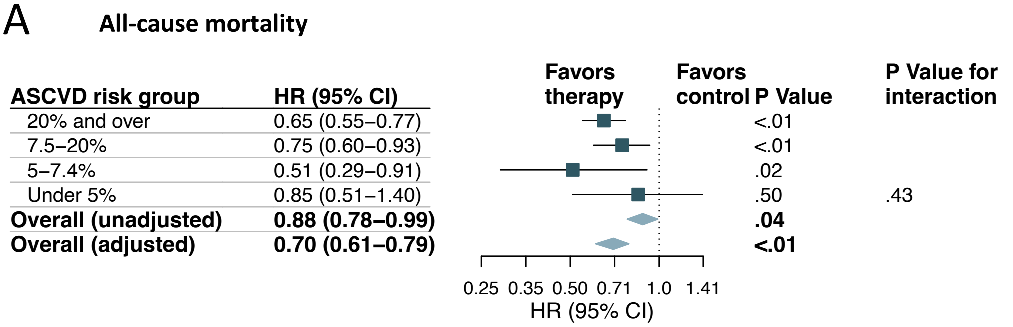


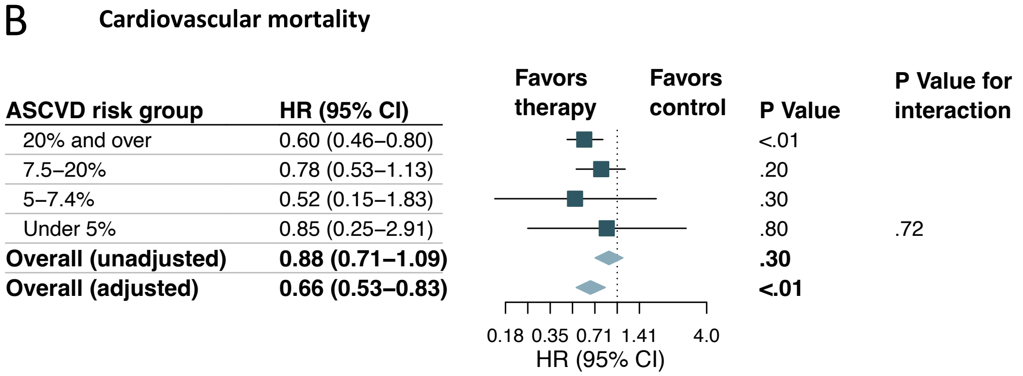


Supplementary figure 5. Analysis of mortality outcomes in the unmatched cohort. (A) Hazard ratios and 95% confidence intervals are shown for all-cause mortality in the total cohort and subgroups with increasing categories of 10-year ASCVD risk. (B) Hazard ratios and 95% confidence intervals are shown for cardiovascular mortality in the total cohort and subgroups with increasing categories of 10-year ASCVD risk.

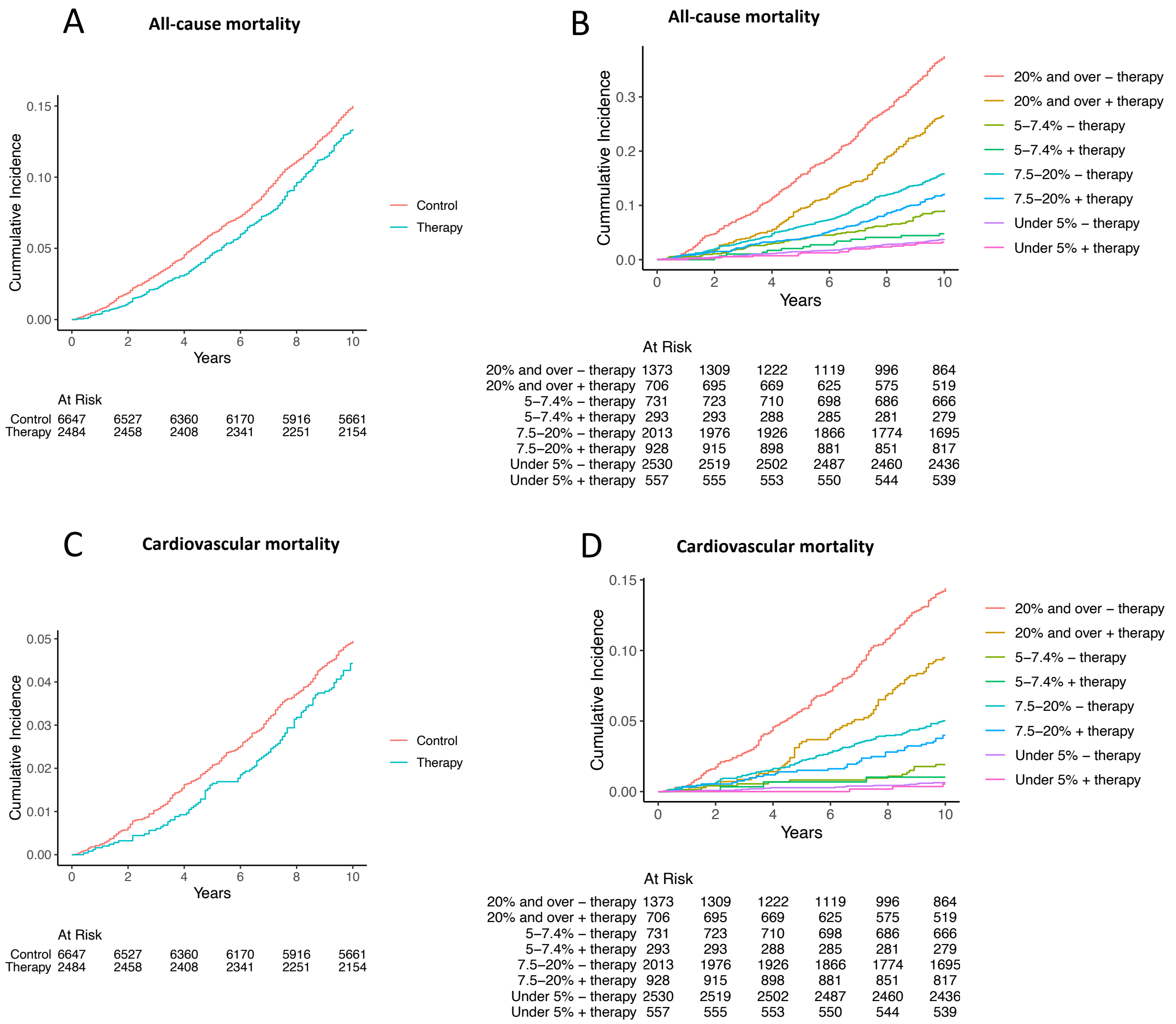


Supplementary figure 6. Cumulative event rate of primary and secondary outcomes in the unmatched cohort. (A) All-cause mortality in individuals who were receiving lipid-lowering therapy, compared to those who were not. (B) All-cause mortality in subgroups with increasing categories of 10-year ASCVD risk. (C) Cardiovascular mortality in individuals who were receiving lipid-lowering therapy, compared to those who were not. (D) Cardiovascular mortality in subgroups with increasing categories of 10-year ASCVD risk.

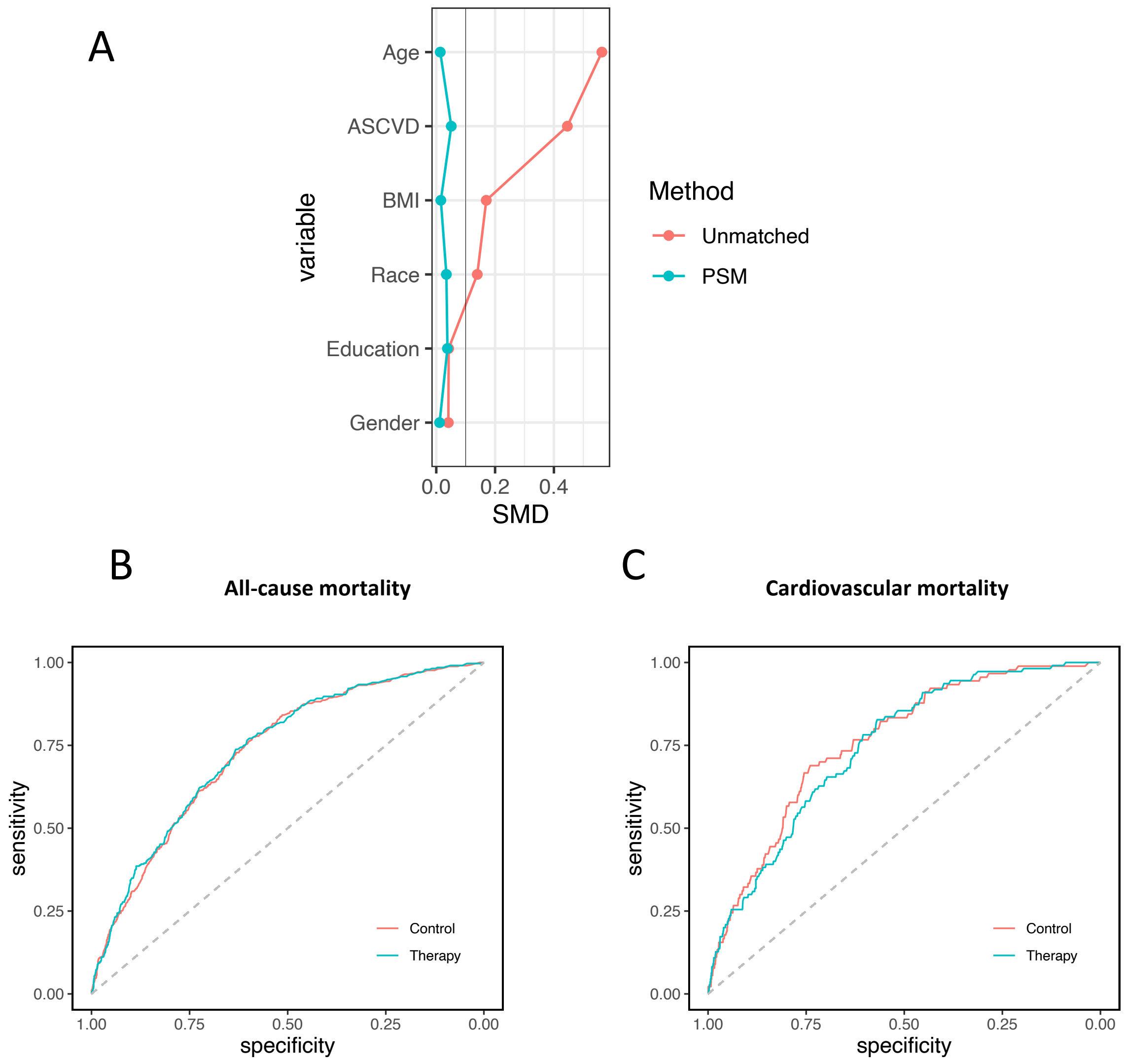


Supplementary figure 7. ROC analysis of 10-year ASCVD risk to predict mortality in the matched cohort using individuals from NHANES 1999-2008 without lipid-lowering therapy as control. (A) Changes in standardized mean difference before and after matching. (B) Predicting all-cause mortality in untreated control and lipid-lowering therapy groups (AUC [95% CI]: 0.73 [0.71-0.76] versus 0.74 [ 0.71-0.76]; P = .80). (C) Predicting cardiovascular mortality in untreated control and lipid-lowering therapy groups (AUC [95% CI]: 0.76 [0.71-0.81] versus 0.75 [0.70-0.79]; P = .67). PSM: propensity score matching.

### A All-cause mortality

| ASCVD risk group | HR (95% CI) | P Value | P Value for interaction |
|---|---|---|---|
| 20% and over | 0.78 (0.64–0.94) | .01 | |
| 7.5–20% | 0.65 (0.51–0.83) | <.01 | |
| 5–7.4% | 0.65 (0.34–1.24) | .20 | |
| Under 5% | 0.74 (0.42–1.34) | .30 | .72 |
| **Overall (unadjusted)** | **0.76 (0.66–0.88)** | **<.01** | |
| **Overall (adjusted)** | **0.77 (0.66–0.89)** | **<.01** | |

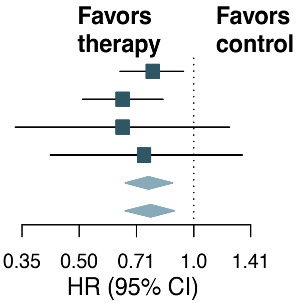


### B Cardiovascular mortality

| ASCVD risk group | HR (95% CI) | P Value | P Value for interaction |
|---|---|---|---|
| 20% and over | 1.00 (0.71–1.41) | >.90 | |
| 7.5–20% | 1.53 (0.89–2.61) | .12 | |
| 5–7.4% | NA | NA | |
| Under 5% | 0.93 (0.21–4.15) | >.90 | .60 |
| **Overall (unadjusted)** | **1.19 (0.90–1.57)** | **.20** | |
| **Overall (adjusted)** | **1.17 (0.88–1.56)** | **.30** | |

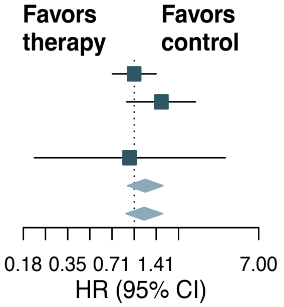


Supplementary figure 8. Analysis of mortality outcomes in the matched cohort using individuals from NHANES 1999-2008 without lipid-lowering therapy as control. (A) Hazard ratios and 95% confidence intervals are shown for all-cause mortality in the total cohort and subgroups with increasing categories of 10-year ASCVD risk. (B) Hazard ratios and 95% confidence intervals are shown for cardiovascular mortality in the total cohort and subgroups with increasing categories of 10-year ASCVD risk.

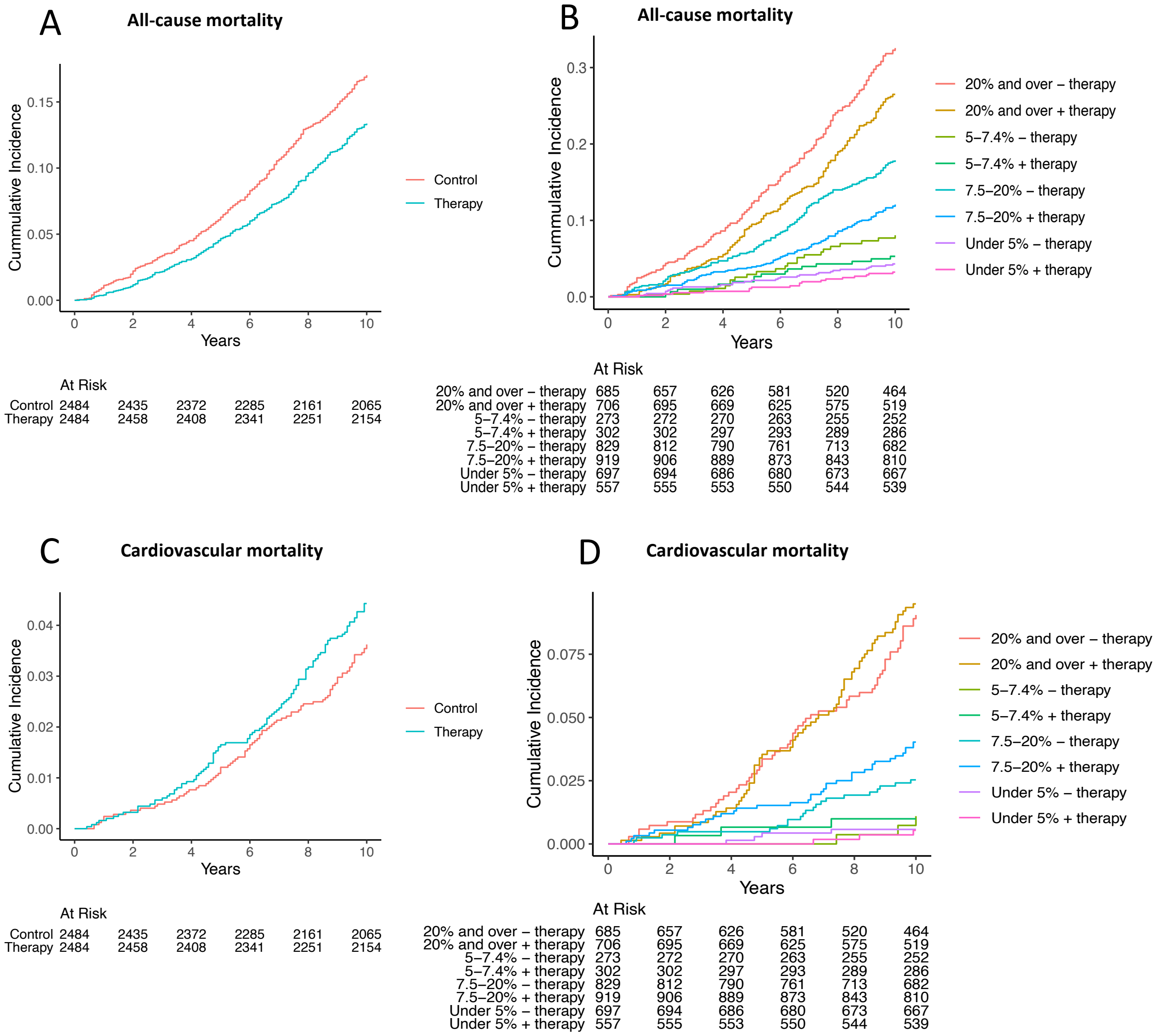


Supplementary figure 9. Cumulative event rate of primary and secondary outcomes in the matched cohort using individuals from NHANES 1999-2008 without lipid-lowering therapy as control. (A) All-cause mortality in individuals who were receiving lipid-lowering therapy, compared to those who were not. (B) All-cause mortality in subgroups with increasing categories of 10-year ASCVD risk. (C) Cardiovascular mortality in individuals who were receiving lipid-lowering therapy, compared to those who were not. (D) Cardiovascular mortality in subgroups with increasing categories of 10-year ASCVD risk.

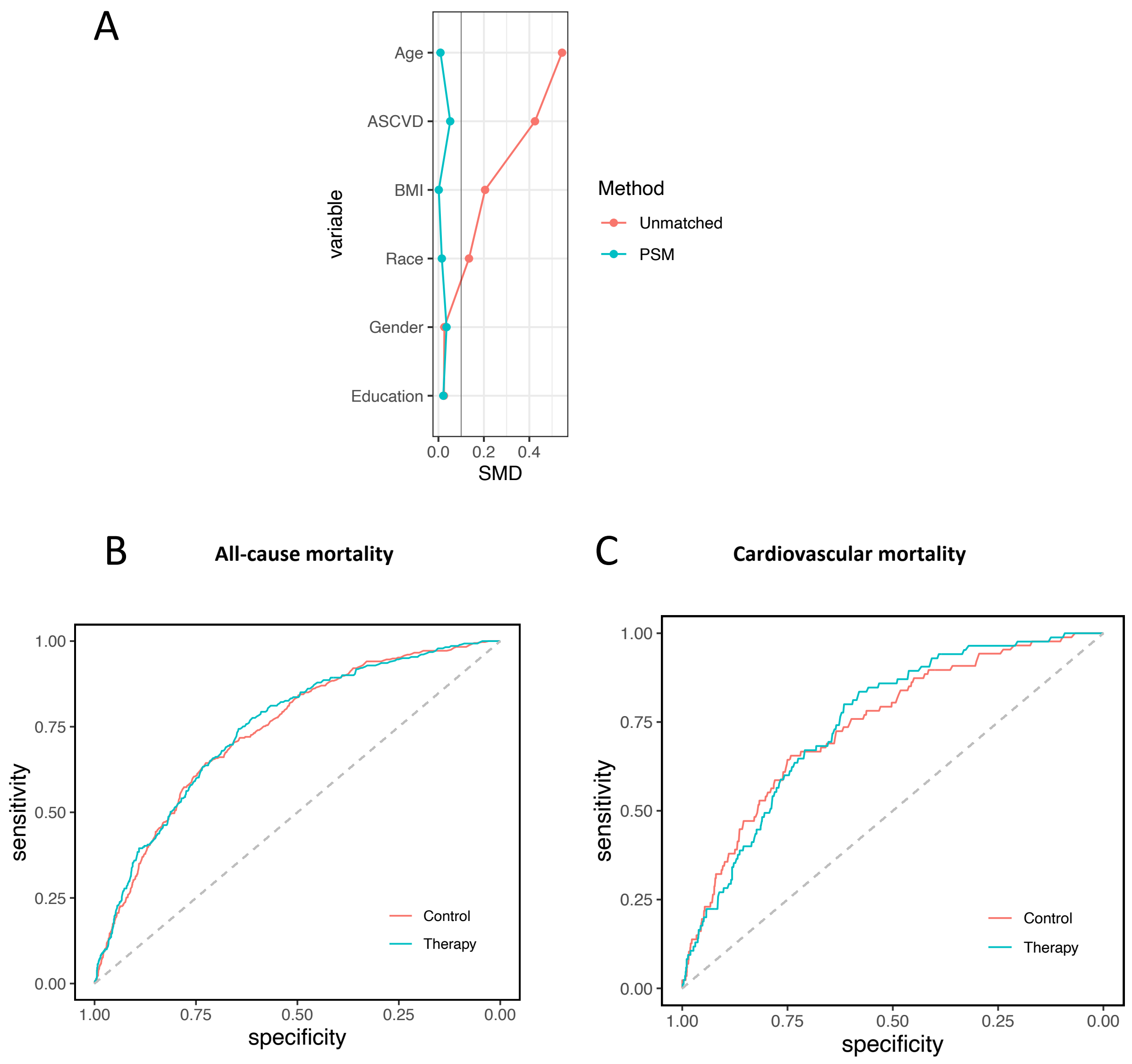


Supplementary figure 10. ROC analysis of 10-year ASCVD risk to predict mortality in the matched cohort from NHANES 1999-2008. (A) Changes in standardized mean difference before and after matching. (B) Predicting all-cause mortality in untreated control and lipid-lowering therapy groups (AUC [95% CI]: 0.74 [0.71-0.77] versus 0.75 [ 0.72-0.78]; P = .73). (C) Predicting cardiovascular mortality in untreated control and lipid-lowering therapy groups (AUC [95% CI]: 0.74 [0.69-0.81] versus 0.75 [0.70-0.80]; P = .82). PSM: propensity score matching.

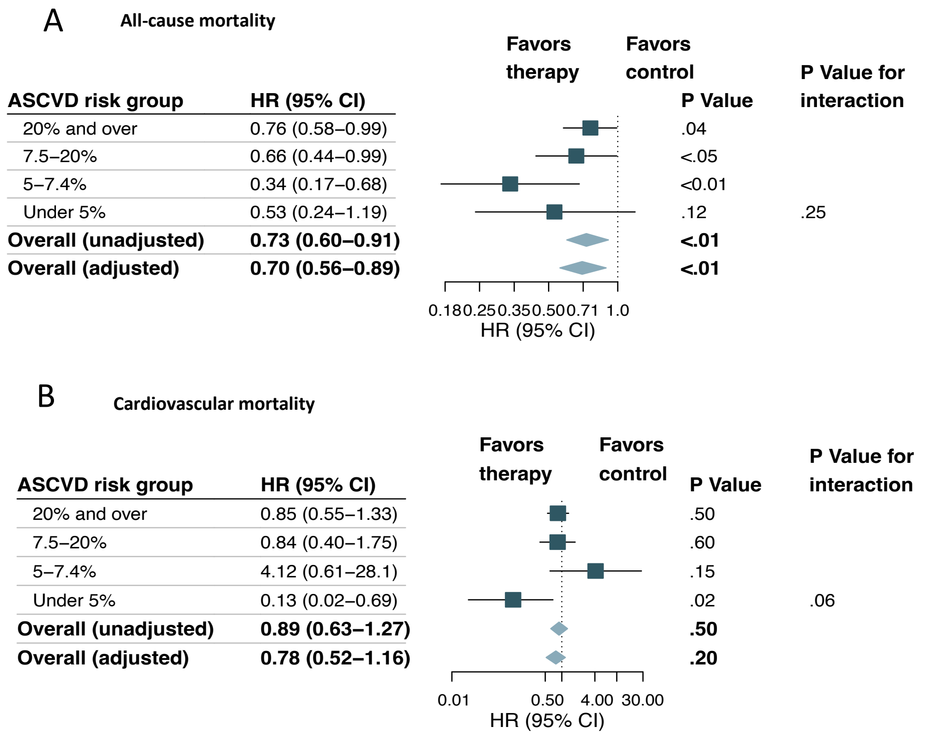


Supplementary figure 11. Analysis of mortality outcomes in the matched cohort from NHANES 1999-2008 using weighted Cox regression. (A) Hazard ratios and 95% confidence intervals are shown for all-cause mortality in the total cohort and subgroups with increasing categories of 10-year ASCVD risk. (B) Hazard ratios and 95% confidence intervals are shown for cardiovascular mortality in the total cohort and subgroups with increasing categories of 10-year ASCVD risk.

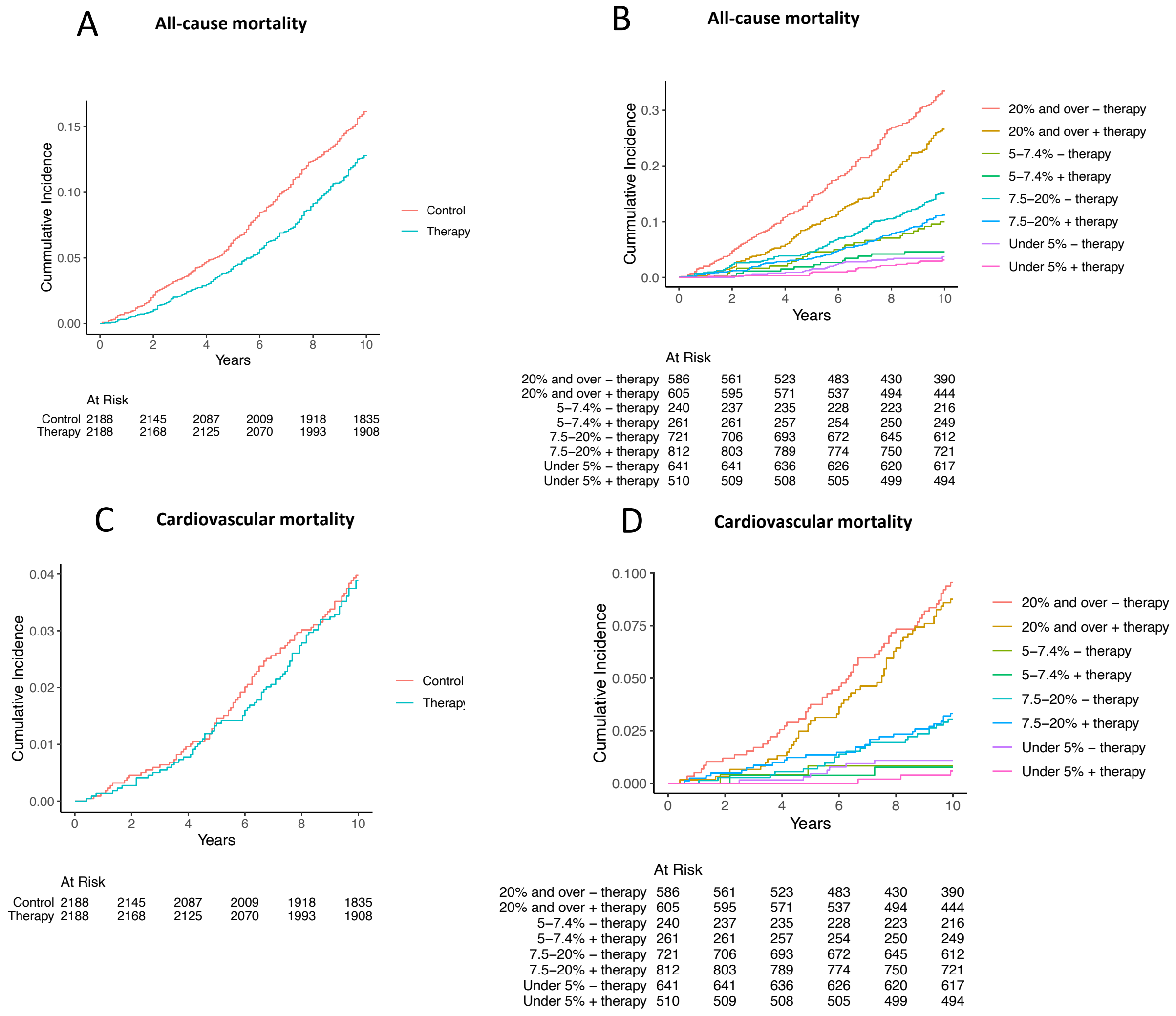


Supplementary figure 12. Cumulative event rate of primary and secondary outcomes in the matched cohort from NHANES 1999-2008. (A) All-cause mortality in individuals who were receiving lipid-lowering therapy, compared to those who were not. (B) All-cause mortality in subgroups with increasing categories of 10-year ASCVD risk. (C) Cardiovascular mortality in individuals who were receiving lipid-lowering therapy, compared to those who were not. (D) Cardiovascular mortality in subgroups with increasing categories of 10-year ASCVD risk.
